# Impact of AI-based Real Time Image Quality Feedback for Chest Radiographs in the Clinical Routine

**DOI:** 10.1101/2021.06.10.21258326

**Authors:** Joerg Poggenborg, Andre Yaroshenko, Nataly Wieberneit, Tim Harder, Axel Gossmann

## Abstract

**Purpose:** To implement a tool for real time image quality feedback for chest radiographs into the clinical routine and to evaluate the effect of the system on the image quality of the acquired radiographs.

**Materials and Methods:** A real time Artificial Intelligence (AI) image quality feedback tool is developed that analyzes chest PA x-rays right after the completion of the examination at the x-ray system and provides visual feedback to the system operator with respect to adherence to desired standards of collimation, patient rotation and inspiration. In order to track image quality changes over time, results were compared to image quality assessment for images, acquired prior to system implementation.

**Results:** Compared to the image quality prior to the installation of the real time image quality feedback solution, it is shown that a relative increase of images with optimal image quality with respect to collimation, patient rotation and inspiration is achieved by 30% (p<0.01). A relative improvement of 28% (p<0.01) is observed for the increase of images with optimal collimation, followed by a relative increase of 4% (p<0.01) of images with optimal inspiration. Finally, a detailed analysis is presented that shows that the average unnecessarily exposed area is reduced by 34% (p<0.01).

**Discussion:** This study shows that it is possible to significantly improve image quality using a real time AI-based image quality feedback tool. The developed tool not only provides objective and impartial criticism and helps x-ray operators identify areas for improvement, but also gives positive feedback.

**Key Findings:**

- A substantial amount of images acquired in the clinical routine does not suffice the international guidelines
- Continuous AI-based image quality feedback to the x-ray system operator in the clinical routine leads to a significant image quality improvement over time
- Using the developed tool, the overall fraction of images with optimal patient positioning could be improved by 30%, followed by a 34% decrease of unnecessarily exposed area.

## Introduction

Optimal patient positioning and image quality are of essential character for proper diagnostic image evaluation and subsequent clinical decision making (1,2). At the same time, insufficient positioning precision may lead to time-consuming recalls, missed diagnoses and increased patient dose through retakes. Moreover, improper positioning may lead to drop in referrals (3). Precise patient positioning for imaging requires adequate positioning time and operator skill. Simultaneously, during the past years the clinical community has been facing an ever-increasing pressure to increase productivity while lowering the total cost (4,5). One of the implications thereof has been a trend to accept image quality that just allows image interpretation in the clinical routine (6), especially for such high-volume imaging modalities as x-ray. Thus, previous reader-based studies have reported substantial deviation from international guidelines (7,8) for chest radiographs acquired in the clinical routine, revealing that less than 50% (9) or even as low as 4% (10) of the images satisfied guideline criteria. Other research highlighted the necessity for continuous image quality control (11,12) in order to ensure proper image quality. The most common finding of the past studies revealed that collimation is often not performed according to the international guidelines (13–16) and the main challenge is operator skill (17). Since the introduction of Picture Archiving and Communication System (PACS) the interaction and communication between radiographers and radiologists have been significantly reduced (18), culminating in tele-radiological services. This trend has intensified the high variability of image quality perception observed both between radiologists (19) and especially between radiologists and radiographers (20). Thus it could be shown that radiologists agreed only in 60% of cases with the necessity to retake a chest posterior anterior (PA) x-ray radiograph, that had been previously rejected by the radiographers (21). Consequently, a real time tool for automatic and objective image quality control could help standardize, improve and track image quality in the clinical routine.

Recently, a neural-network-based algorithm has been developed that can automatically assess quality of collimation, patient rotation and inspiration status on chest PA x-rays (22). Further studies showed that the algorithm yields analog evaluation of image quality with regard to the mentioned parameters to expert radiologists for real world clinical data acquired at different clinical sites compared to the data used for algorithm training (23,24). It could be also shown that while the algorithm accurately determines physical parameters that are used for image quality analysis, there is only a moderate inter-reader agreement for assessment of image quality between human readers (22,24) which is well in line with previous studies (19–21).

To highlight the clinical necessity, the developed Artificial Intelligence (AI) algorithm was tested as an analysis tool for more than 3000 chest radiographs per site acquired in the clinical routine in four different institutions, located in Europe and the US (23,24). In these unpublished data the percentage of images with suboptimal parameters determined by the algorithms for chest x-rays was significantly higher than 50%, which correlates well with previous reports (9,10).

Hence, the goal of the current study was to demonstrate the feasibility of implementing the developed AI-based tool for real time objective and comprehensible feedback to the radiographer after each image acquisition into the clinical routine. The study hypothesis was that through the continuous feedback received by the radiographers the overall image quality would significantly improve.

## Materials and Methods

During this study only images acquired during the normal clinical routine were analyzed post-acquisition with respect to image quality. The study was IRB approved by the local ethics committee.

The algorithmic evaluation of collimation, patient rotation and inspiration status is based on the pretrained neuronal network (22) that detects the lung, the ribs, the clavicles and the backbone. Subsequently the algorithm calculates the minimal distance between the lung and the image border for all four directions separately. In order to assess the precision of the collimation, the calculated distances are compared to the respective thresholds for minimal and maximal clinically desirable collimation, which were determined based on expert radiologist input prior to the study (23,24). Patient rotation is evaluated based on the asymmetry of the clavicles with respect to the center of the backbone, and the number of ribs that are covered by the lung are used a figure of merit for estimation of inspiration status. The thresholds that were used for the respective parameter to distinguish between precise and imprecise positioning in the study are summarized in Table 1. An x-ray of the chest was considered as optimal only if all three analyzed parameters – collimation, patient rotation and inspiration are within the pre-defined thresholds.

**Table 1:**
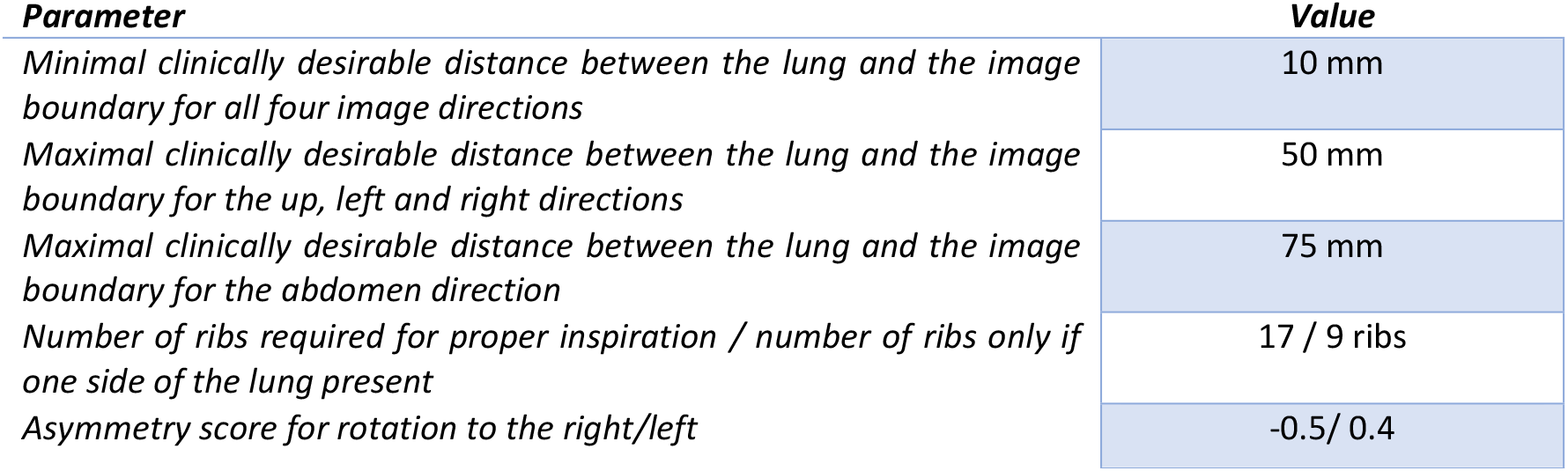
Thresholds used to distinguish between precise and imprecise patient positioning (25).

The described AI algorithm was implemented into the clinical routine using the IT infrastructure sketched out in Figure 2. An x-ray radiography system DigitalDiagnost 3.1 (Philips Medical Systems DMC, Germany) was configured to send the acquired x-ray radiograph to PACS for reading purposes as this is done in the normal clinical routine while a copy of the image was sent to the server with the installed AI image quality evaluation algorithm in parallel. The result of the image quality analysis was then displayed to the user on a tablet PC, located at the acquisition console of the x-ray system (Figure 3). The complete solution, consisting of the AI algorithm and the IT infrastructure is referred to as Smart Assistant.

**Figure 1:**
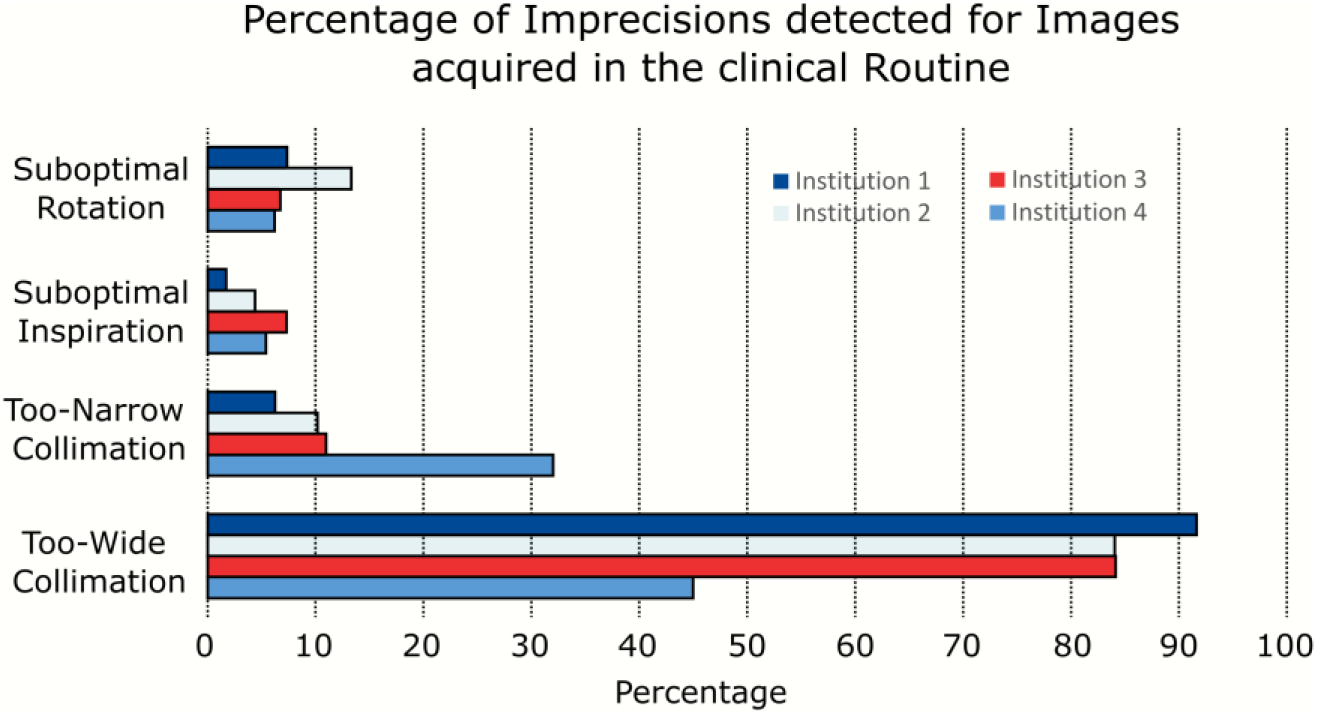
Percentage of chest PA x-rays with patient positioning imprecisions

**Figure 2:**
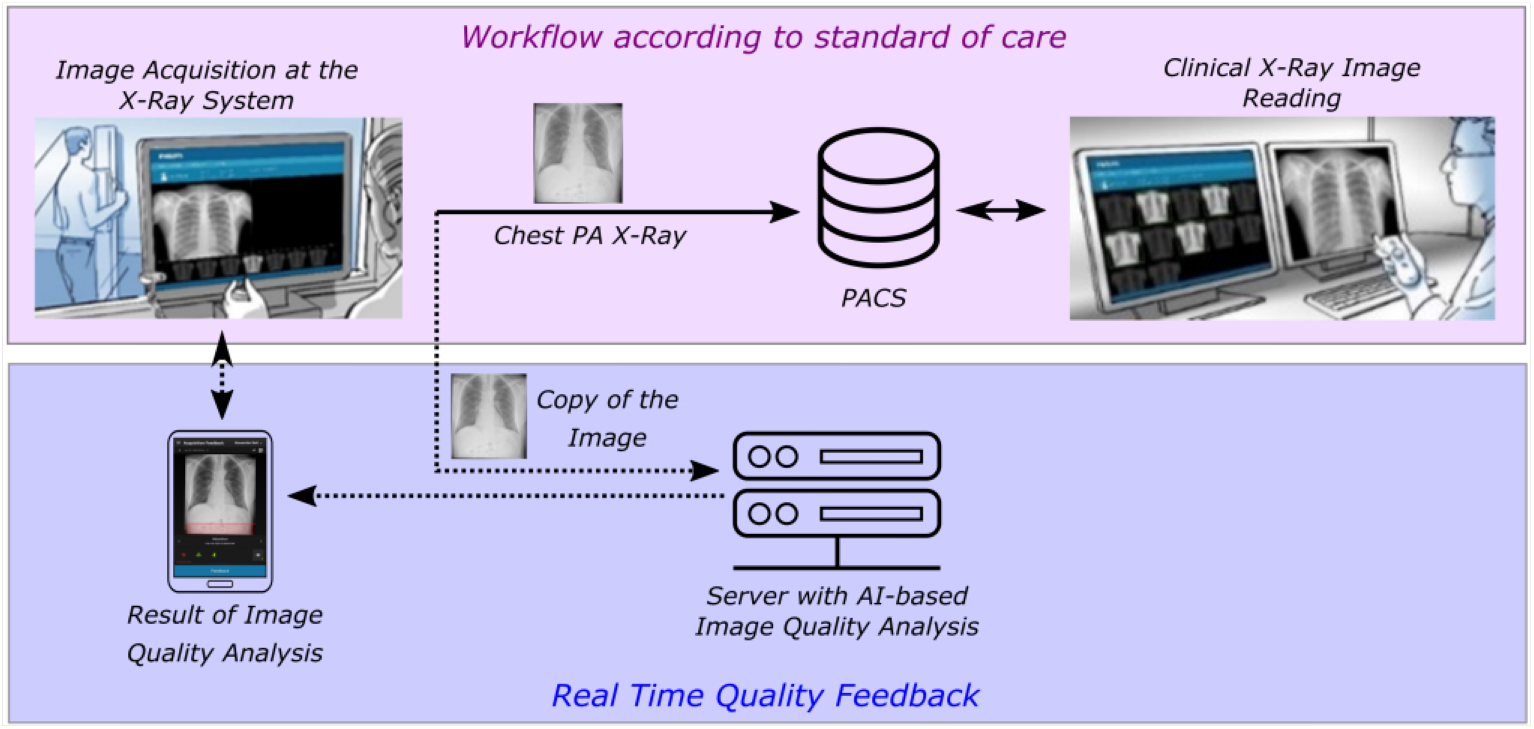
IT infrastructure used for the real time AI feedback. An image acquired in the clinical routine is sent according to the standard of care for diagnostic reporting to PACS. A copy of the image is generated and sent to a separate server with the AI software. The result of image quality analysis is displayed on a tablet PC, located at the x-ray system console.

**Figure 3:**
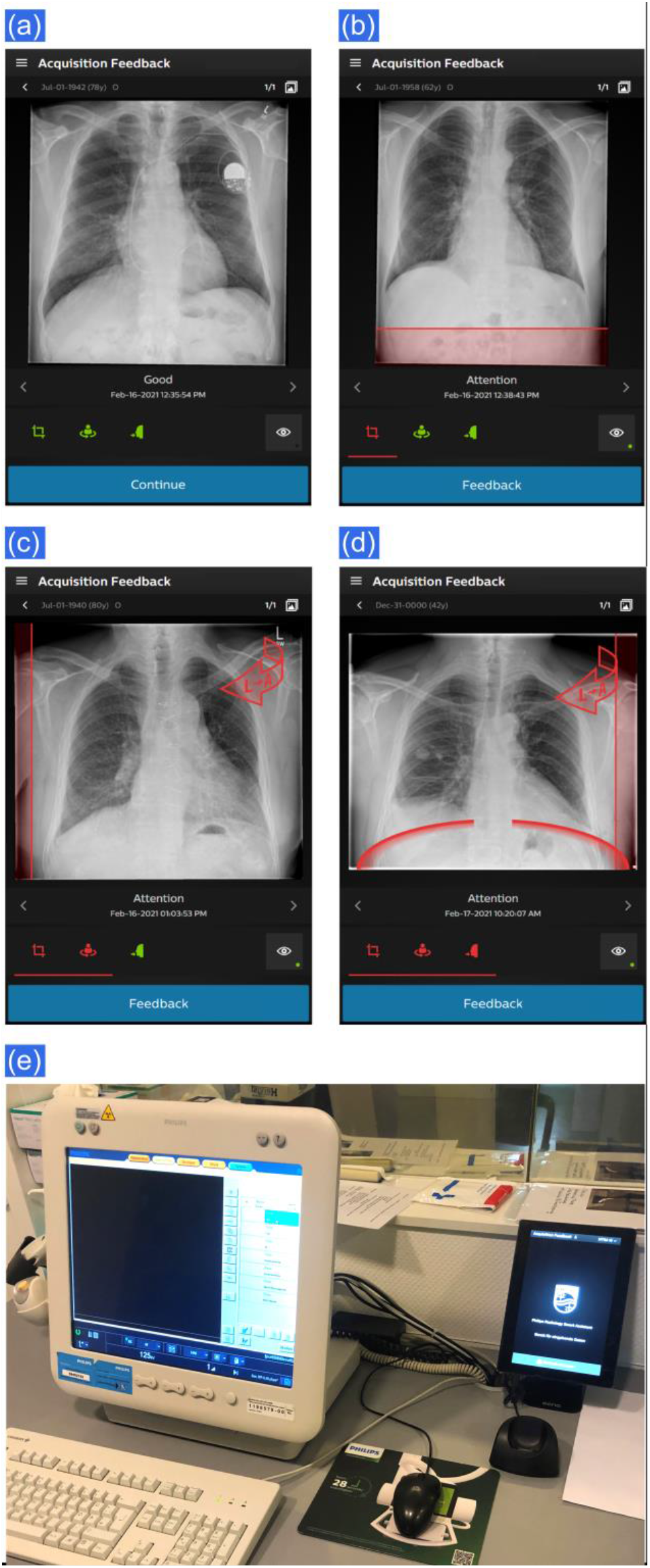
Sample of visual feedback displayed on the tablet PC for: optimal image quality (a); image with collimation that is too wide in the abdomen region (b); image with a subotimal patient rotation and too wide collimation (c); image with insufficient inspiration, patient misrotation and too wide collimation (d). Right: a photograph of the workstation and the corrsponding tablet PC (e).

Based on the result of the algorithmic analysis, the image quality parameters below the image are highlighted in green on the tablet PC, if the image quality is considered to be optimal. In case the collimation is too wide, the unnecessarily exposed area is marked with a red overlay. In case of collimation that is too narrow, the corresponding side(s) are marked by a red bar (Figure 3 (a)-(d)).

The developed Smart Assistant prototype was evaluated in the clinical study that was performed as follows: the retrospective image quality analysis was performed before the system was implemented and the active clinical phase started after implementation of the system. For the retrospective analysis, chest PA radiographs acquired in the clinical routine prior to the initiation of the clinical study were retrieved from PACS and analyzed using the developed image quality algorithm. For the retrospective analysis and the active phase, 2774 images acquired during 12 months of clinical routine and 1973 images imaged during 11 months, respectively, were analyzed. Based on previous studies (22–24), no additional validation of the algorithmic performance was performed. Only images acquired with the same x-ray system as later used for the active phase were considered for the retrospective analysis. The results of the retrospective positioning quality analysis were documented for later comparison with data obtained during the active phase.

During the active phase of the study, Smart Assistant for real time AI-based image quality feedback was installed in the clinical routine and provided feedback to the x-ray system operators after each image acquisition. The workflow consisted of the following steps: a chest radiograph is acquired and the radiographer finalizes image acquisition on the x-ray modality and sends the image to PACS. A copy of the image is simultaneously sent to a server on which the AI image quality evaluation package is installed (see Figure 2). The image is analyzed and the feedback is presented to the radiographer on the tablet PC. In case the algorithm determined image quality to be optimal the radiographer receives a respective feedback and there is no further need for the user to interact with the tablet PC. In case, however, one or more of the parameters are found to be outside the predefined optimal range, the parameter is marked in red, and the operator has to interact with the tablet PC to acknowledge the feedback and to select a reason from a pre-defined list what he or she believes to be the root cause of the imprecision. The list consists of 6 possible pre-selected reasons, for example: “bariatric patient”, “language barrier”, etc. The result of each image quality analysis is saved in a log-file that is used to track the image quality statistics. No image retakes were performed by the radiographers based on the feedback of the AI-algorithm and the process for retakes relied on the standard clinical routine. In cases, when a retake of the x-ray was considered, a radiologist took a decision after inspecting the image. The team performing image acquisition for the retrospective and the active phase consisted of 15 radiographers with varying experience in chest radiography imaging.

Based on the documented output of the algorithmic analysis, the imaged area outside the maximum clinically desirable collimation thresholds and the area cut off with respect to the minimum clinically desirable collimation thresholds was calculated for each image. The two areas will be referred to as unnecessarily exposed and cut-off areas in the following. In particular, the calculation of the areas was based on the result of the algorithmic assessment for the minimum distance between the lung and the image border. Subtraction of the maximum/minimum clinically desirable threshold from the derived value provided a distance that has been overexposed/cut-off for each of the four image sides. Multiplication of the corresponding distances resulted in the aforementioned areas. Figure 4 (a) shows an exemplary image, where the minimal and maximum clinically desirable collimation thresholds are marked with the dotted and the dashed lines, respectively. In the shown case, the unnecessarily exposed area is marked with a red overlay. The cut-off area was calculated as the area between the image border and the minimal clinically meaningful collimation threshold. Figure 4 (b) shows a cut-off image, where the cut-off area is highlighted in red.

**Figure 4:**
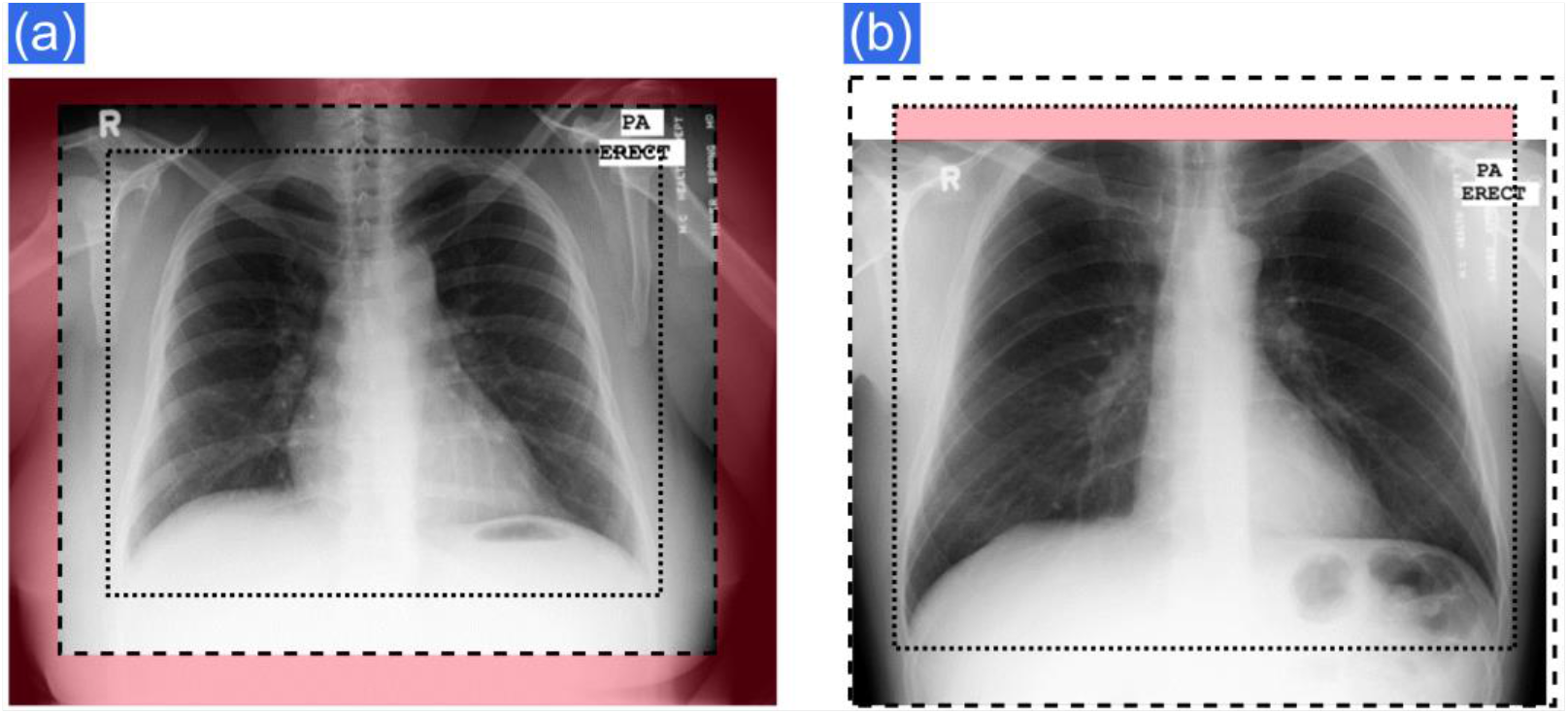
Examples of an image with a collimation that is too wide (a), where the unnecessarily exposed area is marked with a red overlay. The minimal and maximal clinically desirable collimation thresholds are marked with dotted and dashed lines, respectively. On image (b) the area marked with red, corresponds to the cut-off area.

Image quality parameters obtained for the retrospective and the active phases were valuated using Excel 2016 (Microsoft, USA). The parameters for the percentage of images were compared using a Pearson’s chi-squared Test. The unnecessarily exposed and cut-off areas were compared using the Mann-Whitney test. All results were considered to be statistically significant if p<0.05.

## Results

The developed AI-based image quality feedback could be implemented without any problems into the clinical workflow. The time delay between completing an image at the x-ray system and the visual feedback is on the tablet PC was measured to be approx. 10 seconds. This time includes the time required by the system for the image export process, the traffic time to the server, the time for the image quality analysis (less than 1 second per image) and the transmission of the feedback to the tablet. There were no complications for the clinical routine workflow related to the use of the tool.

Compared to the retrospective baseline analysis, a clear improvement in image quality parameters was observed during the active phase. The fraction of images with overall optimal positioning showed a significant improvement of 30% during the active phase of the study relative to the retrospective analysis. This improvement was largely driven by a 28% increase in the fraction of images with optimal collimation, followed by a 4% increase of the fraction of images with optimal inspiration. A positive trend, but no statistically significant improvement was observed for the fraction of images with (sub)optimal rotation of the patient. The detailed results of the image quality analysis are summarized in Table 2. The relative change presented in the last column represents the improvement divided by the value obtained for the retrospective analysis.

**Table 2:** Summary of the image quality analysis performed before (retrospective) and after installation (active phase) of the AI solution. The numbers in parenthesis represent the number of images in the corresponding group.

|  | **Retrospective Analysis** | **Active Phase** | **p value** | **Relative Change** |
| --- | --- | --- | --- | --- |
| % of optimal images | 31.4 % (872) | 40.8 % (802) | < 0.01 | 30 % |
| % of images with Optimal collimation | 34.1 % (947) | 43.5 % (859) | < 0.01 | 28 % |
| % of Images with Optimal Inspiration | 94.6 % (2623) | 98 % (1934) | < 0.01 | 4 % |
| % of Images with Optimal Rotation | 93.8 % (2601) | 94.8 % (1870) | 0.14 | - |

A time-resolved evaluation of the image quality revealed a steady improvement of the parameters, as is demonstrated in Figure 5 for collimation. While there are time periods that show expected oscillating image quality performance (blue line), the linear trendline fitted to the data (red dashed line) shows a steady improvement over time with no stagnation perceptible. Consequently, it could be hypothesized that image quality parameters could improve even further if the trial period was longer.

**Figure 5:**
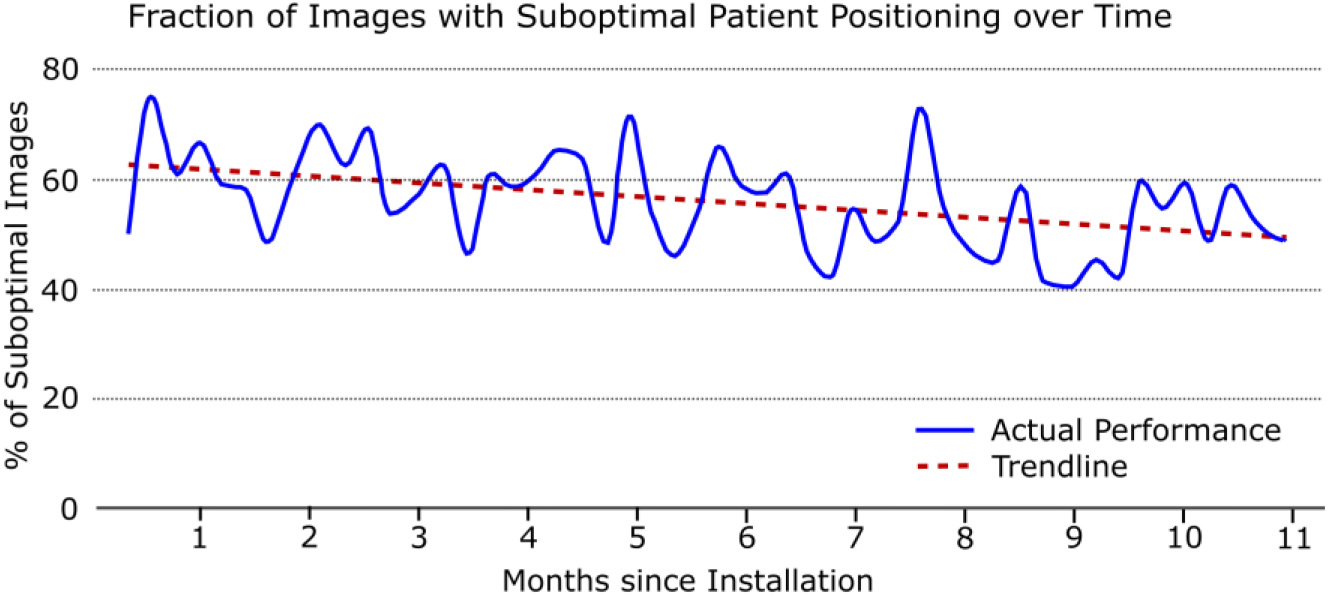
Time-resolved fraction of images with suboptimal patient positioning over the time period of the active phase

A detailed analysis of the collimation issues for the four individual sides of the image revealed, as can be seen in Figure 6 that during the active phase a significant improvement was achieved for both under and over collimation for the top side and for the too-wide collimation on the bottom side. This result seems to be reasonable, given the fact that these are the sides that are most challenging for the radiographers. A further statistically significant change observed was a reduction of images collimated too widely on the left side, which seems to be rather surprising at the first. However, in the given examination room, the radiographer typically stands on the right side, which can be an explanation for this unexpected result.

**Figure 6:**
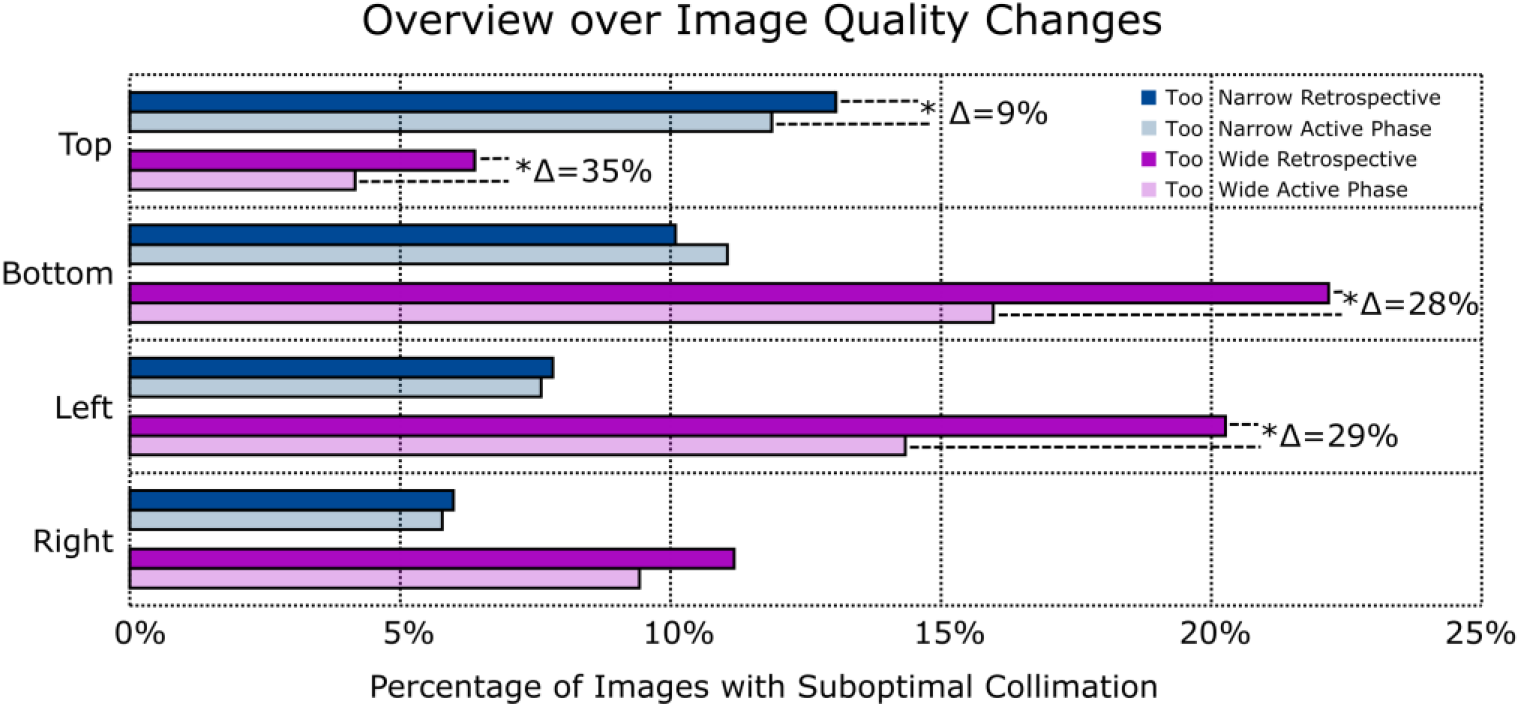
Percentage of images with the collimation issues broken up for the four different sides of the image for the retrospective analysis and the active phase. Statistically significant changes with p<0,05 are marked with *.

Collimation imprecisions result in unnecessarily exposed as well as cut-off areas. Both parameters were calculated for all images separately and it was observed that the average unnecessarily exposed area, i.e. the area outside the maximum clinically desirable collimation threshold could be reduced during the active phase by 34% (see Table 3) compared to the retrospective analysis. At the same time, a positive trend, but no significant change, could be observed for the cut-off area, i.e. the area that is lacking to achieve the minimum clinically desirable collimation.

**Table 3:** Comparison of the unnecessarily exposed and cut-off areas for the retrospective analysis and the active phase.

|  | Retrospective Analysis | Active Phase | p value | Relative Change |
| --- | --- | --- | --- | --- |
| Average Unnecessarily Exposed Area | 31.0 cm<sup>2</sup> | 20.1 cm<sup>2</sup> | < 0.01 | 34 % |
| Average Cut-Off Area | 9.7 cm<sup>2</sup> | 9.5 cm<sup>2</sup> | 0.41 | - |

Thus, the presented results clearly demonstrate that the image quality significantly improved during the active phase of the AI-based image quality feedback after the tool was implemented into the clinical workflow. Especially, the too-wide collimation was significantly reduced, while also a positive trend was seen for the too-narrow collimation.

## Discussion

In this study, we have implemented a system for a fully automated real time AI-based image quality feedback for PA chest radiographs into the clinical routine workflow. The majority of the approx. 10 second delay time between the finalization of the study at the x-ray system and the feedback is related to the time the x-ray system needs for the image export process and the traffic time, as the image analysis itself takes less than 1 second. The delay time was estimated during the normal clinical routine, however, it is pointed out that it is influenced by the network load and the number of images in the examination. In the future, an optimization of the IT infrastructure, such as prioritization of the traffic to the AI server or direct connection of the server to the x-ray system could be key in order to further reduce the time delay and make the workflow even smoother.

The key finding of the study is that the developed tool helps to improve image quality in the clinical routine. The greatest impact of the tool was observed in the reduction of the rate of images that displayed a collimation that was to wide, an issue that has been identified as a significant deviation from clinical guidelines by previous studies (13–16). This improvement associated with a significant reduction of the average unnecessarily exposed area during image acquisition. Thus, the achieved improvements help to reduce patient dose, but they will also contribute to reduce scattered radiation (26–29). The optimal collimation in left-right direction also suggests proper centering of the patient in front of the automatic exposure chambers. At the same time, a positive trend, but no statistical improvement for the average cut-off area could be observed. There are several reasons why there was no statistical improvement for the cut-off area. First, the average cut-off area is very small in the first place. The threshold for minimum collimation for all four image directions was set at 10 mm for the study. If one now considers that the average collimation opening is approx. 30 cm (corresponding to the size of an adult thorax), then the average cut-off area of 9,5 cm^2^ corresponds to a collimation deviation of 3,2 mm in one direction. Therefore, on average the whole of the lung is present on the image and only the distance between the lung and the image border is smaller than the pre-determined threshold. As a further outcome of the study, a reduction of images with suboptimal inspiration was observed. Evaluation of chest x-rays with suboptimal inspiration often complicates diagnosis, as the reader has no indication whether improper inspiration is related to a clinical condition or imprecise image acquisition. Therefore, a reduction of images with insufficient inspiration may improve the diagnostic process and therefore also has positive impact on patient outcome.

Image quality is directly linked to the efficiency of the clinical workflow (1,3). As a consequence, there is a high probability that costs will decrease through usage of the developed tool. Optimal image quality increases the confidence during clinical image reading (30), prevents recalls and reduces the probability for a follow-up imaging (31). Only few studies in the past have sought to quantitatively analyze the relationship between image quality and workflow efficiency. Siegel at al. (30) have reported that an increase of 16% in reading time was required by radiologists for x-ray chest radiographs with suboptimal image quality, while another study suggested that the effect of suboptimal image presentation strongly depends on the reader experience (32).

An aspect that should not be overlooked is that the developed tool introduces a possibility for the radiographers to objectively track their performance and help them to identify areas for personal improvement. Positive feedback received for optimal patient positioning can be a great motivational aspect, as has been pointed out by previous studies (33).

One of the limitations of the described study is that the results were obtained during the onset of the Covid-19 pandemic, which implied that no additional training/workshops could be provided to the technologists during the active phase. It can be hypothesized that additional guidance on patient positioning for issues identified with the developed tool from experienced quality managers would improve the results even further. Additionally, the improvement over time did not show any signs of curve flattening so that a further improvement can be expected if the tool was used for a longer period of time.

A further limitation of the present study is that it does not take into account the additional clinical information, such as intentionally wide collimation e.g. for catheter imaging. Achievable image quality depends further on the patient population imaged in this study and their ability to cooperate with the technologist. This should be considered when discussing the obtained absolute image quality statistics. In order to account for this and to make image quality changes generalizable, the chosen time periods for the retrospective and the active phase were of comparable length and the number of images for the two phases was also similar.

In summary, the presented study shows that AI-based real time image quality feedback can be used in the clinical routine to significantly improve image quality. The developed tool holds high potential for transferring the method to further anatomies.

## Summary

The impact of the developed AI-based real time image quality feedback tool was evaluated in the clinical routine, showing that the real time, standardized, objective image quality feedback for the x-ray system operator leads to a significant increase of image quality over time.

## Data Availability

Image quality statistics obtained during this study are available upon legitimate request

## Acknowledgments

The authors would like to thank U. Galano, C. Mueller, I. Brandes, L. Tanski and B. Hoenisch for very fruitful discussions on the design of the user interface, and D. Fahlbusch and J. Eikermann for the input on prototype development.

